# The Probability of Choosing Both Hands Depends on an Interaction Between Motor Capacity and Limb-Specific Control in Chronic Stroke

**DOI:** 10.1101/2020.05.20.20104299

**Authors:** Rini Varghese, Jason J. Kutch, Nicolas Schweighofer, Carolee J. Winstein

## Abstract

A goal of rehabilitation after stroke is to promote pre-stroke levels of arm use for everyday, frequently bimanual, functional activities. We reasoned that, after a stroke, the choice to use one or both hands for bimanual tasks might depend not only on residual motor capacity but also the specialized demands imposed by the task on the paretic hand. To capture spontaneous, task-specific choices, we covertly observed 50 pre-stroke right-handed chronic stroke survivors (25 left hemisphere damage, LHD) and recorded their hand use strategies for two pairs of bimanual tasks with distinct demands: one with greater precision requirements (photo-album tasks), and another with greater stabilization requirements (letter-envelope tasks). The primary outcome was the choice to use one or both hands. Logistic regression was used to test the hypothesis that the probability of choosing a bimanual strategy would be greater in those with less severe motor impairment and those with LHD. When collapsed across the four subtasks, we found support for this hypothesis. However, notably, the influence of these factors on bimanual choice varied based on task demands. For the photo-album task, the probability of a bimanual strategy was greater for those with LHD compared to RHD, regardless of the degree of motor impairment. For the letter-envelope task, we found a significant interaction between impairment and side of lesion in determining the likelihood of choosing both hands. Therefore, the manner in which side of lesion moderates the effect of impairment on hand use depends on the task.

## Introduction

Every day, we engage in tasks that are bimanual in nature—buttoning a shirt, lifting a large object, or folding a piece of paper—tasks that naturally elicit a choice to use both hands together (Kilbreath and Heard 2005). After a stroke, bilateral hand use is critically reduced (Vega-González and Granat 2005; Rinehart et al. 2009; Michielsen et al. 2012; Bailey et al. 2014, 2015), oftentimes despite adequate sensorimotor capacity. The inability to return to prestroke patterns of bilateral hand use is associated with poor recovery of function (Haaland et al. 2012). For this reason, there has been growing interest in understanding and potentially promoting post-stroke arm use, with one of the foremost challenges being how to best assess use so that it is a close approximation of behavior in the natural environment.

Controlled laboratory environments in which stroke survivors are directed to perform pre-defined tasks (Przybyla et al. 2013; Coelho et al. 2013; Bailey et al. 2014; Yadav et al. 2019) under set instructions or time limits, impose a restriction on free, self-selected choices. Thus, such an approach, although well-controlled, lacks everyday representativeness and generalizability. In recent years the use of real-world remote monitoring devices, such as accelerometers and activity monitors, has gained attention (Vega-González and Granat 2005; Rinehart et al. 2009; Thrane et al. 2011; Michielsen et al. 2012; Bailey et al. 2014, 2015; Rand and Eng 2015; Franck et al. 2019; Yadav et al. 2019). In this approach, movement counts, or frequency, often averaged across epochs of time, serve as a metric of use, and, simultaneous movement counts of both upper limbs serves as a proxy for bilateral use (Sterr et al. 2002; Vega-González and Granat 2005; Uswatte et al. 2006; Bailey et al. 2014, 2015). Although extremely valuable and relevant for providing telehealth services, this approach does not capture the task-specific nature of everyday functional activities in which the two hands do not simply move together but rather cooperate to accomplish the task goal.

Indeed, the task-specific nature of hand use is perhaps best exemplified in bimanual tasks, which entail a natural division of labor between the hands such that each hand assumes a preferred role (Guiard 1987). Consider, for instance, the skilled action of threading a needle: for a majority of right-handed individuals, the left hand assumes the role of stabilizing the needle, while the right hand passes the thread through its eye.

By studying hand selection patterns in able-bodied adults, some studies have suggested that hand choice results from an interaction of task demands with lateralized motor control processes (Mamolo et al. 2004, 2006; Przybyla et al. 2013; Stone et al. 2013; Coelho et al. 2013). According to one theoretical framework of motor lateralization (the Dynamic Dominance hypothesis), the left hand (primarily under control of the right hemisphere) is more proficient at stabilization of position through impedance control mechanisms; whereas the right hand (primarily under the control of the left hemisphere) is better developed for producing precise movement trajectories through predictive control mechanisms (Sainburg 2002). In the needle-threading example, given that holding the needle in place is an integral part of the task goal and is well-aligned with the presumed competency of the left hand at position stabilization, it follows that the left hand is “selected” (likely implicitly) by the nervous system to fill this role. Conversely, the right hand, adept at precise visuomotor control of the thread’s trajectory, is selected for the equally important complementary goal of passing it through the needle’s eye. This example also illustrates that bimanual tasks, by design, lend themselves for evaluation of the specialized use of the two hands in a functional context. The interaction between task demands and lateralized motor control and their influence on spontaneous choice in chronic stroke survivors, however, has not been formally explored in the context of bimanual tasks.

Taken together, the choice to use one or both hands in the real-world seems to not only be self-selected but also task specific. This observational investigation is motivated by one broad question: In chronic stroke survivors, what factors influence the spontaneous selection of both hands for bimanual tasks— tasks that would otherwise, in age-similar able-bodied individuals, naturally elicit the use of both hands? Based on the previously well-studied influence of motor impairment and side of stroke lesion (Rinehart et al. 2009; Yadav et al. 2019), we tested an a-priori hypothesis that those with more severe motor impairments and those with right (nondominant) hemisphere damage (RHD) would be less likely to choose both hands together.

In particular, given the known lateralization of control discussed above, we posited that the choice of motor strategy would be influenced by task demands, such as the need for stabilization with the left hand, or for precise visuomotor mapping and trajectory control with the right hand. To examine task-specific effects, we selected two pairs of bimanual tasks with distinct requirements: the first pair involved folding a letter and inserting it into an envelope (letter-envelope tasks) and the second involved receiving a large and heavy photo album and inserting a photo into one of the album’s sleeves (photo-album tasks).

We reasoned that the stabilization of lightweight (paper) objects inherent in the letter-envelope tasks warrants the involvement of the left hand, and so might pose greater demands in those with RHD in whom the left hand is weaker. If this is true, in those with RHD, we would expect to see a sharp rise in the selection of both hands for those with less severe motor impairment. Conversely, owing to its weight, receiving the photo album would require a fair amount of strength, but would be self-stabilizing once placed on the table. The need for precise insertion of the photo into the sleeve stipulates greater involvement of the right hand and might impose greater demands on the paretic right hand of those with LHD. Thus, we would expect an increased selection of both hands in LHD for the photo-album tasks.

We validated the bimanual nature of these four tasks in age-similar able-bodied adults. In a secondary analysis, we quantified the time taken to complete each of the four tasks. We reasoned that even though no time limits were imposed on task performance, individuals would likely choose a motor strategy that would represent the most temporally efficient strategy to complete the task. If this hypothesis is correct, for the stroke group, we would expect there to be no difference in movement time between those who choose a unimanual strategy compared to those who choose a bimanual strategy for any given task.

## Methods

### Participants

Fifty pre-stroke right-handed chronic stroke survivors (25 left hemisphere damage, LHD) and 11 age-similar able-bodied controls gave informed consent to participate. Of the 50 stroke survivors, 42 were enrolled as part of a larger phase-IIb clinical trial (Dose Optimization for Stroke Evaluation, ClinicalTrials.gov ID: NCT01749358, Winstein et al. 2019), whereas 8 were recruited as part of a pilot study in collaboration with the Moss Rehabilitation Research Institute (Einstein Healthcare Network, PA). Study protocol and informed consent was approved by the Institutional Review Boards for the Health Sciences Campus of the University of Southern California and the Einstein Healthcare Network and was in accordance with the 1964 Declaration of Helsinki.

### Motor Component of the Upper Extremity Fugl-Meyer (UEFM)

The motor component of the UEFM is a measure of impairment of the contralesional arm and hand after stroke and involves tests of strength and independent joint control. Item-wise scoring of the UEFM ranges from 0 (unable to perform) to 2 (able to perform completely) while total score ranges from 0 to 66, with a higher score indicating lesser impairment (Fugl Meyer et al. 1975).

### Assessment of Choice

Spontaneous choice was assessed using items of the original Actual Amount of Use Test (AAUT). (Taub et al. 1998) The AAUT is designed to assess spontaneous upper limb use in stroke survivors for a series of seventeen tasks (3 postural items and 14 task-specific items, see Supplementary Material I). The unique aspect of the AAUT is its covert administration; participants who have given prior consent to be video-taped, perform the task battery without any instructions, supervision, or time limits, and performance is captured on video unbeknownst to the participants, who are later debriefed at the end of the assessment (see Supplementary Material I for more information on how the covert nature of the test is operationalized). The open-ended nature of the AAUT allows an objective assessment of spontaneous unobtrusive arm use behavior in a quasi-naturalistic setting with ecologically valid tasks. Video data were recorded at 30 fps and were available for offline observational analysis.

We selected four of the seventeen items based on an a-priori assumption that these items would naturally elicit bimanual use in able-bodied adults. We validated this assumption by collecting data in age-similar able-bodied adults to confirm that these tasks did indeed elicit a bimanual strategy. The four items were: (1) Fold Letter (standard US Letter, 8.5 × 11”), (2) Insert Letter into Envelope (standard commercial envelope, size 9, approx. 4 × 9”), (3) Receive Photo Album (standard 3-ring binder, 2 × 9 × 12”, 2.5 lbs.) and (4) Insert Photo in Album Sleeve (4 × 3.5” polaroid images into a clear sleeve with two 4 × 6” pockets). Fig. 1 illustrates the 4 subtasks.

**Figure 1:**
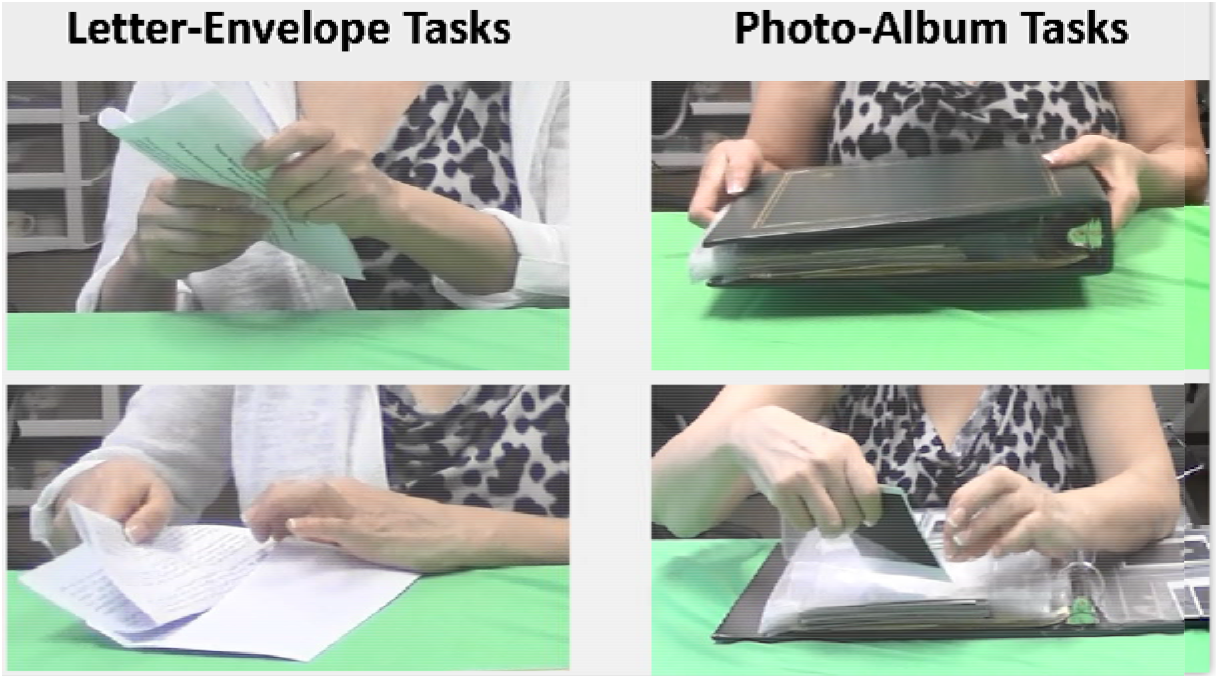
*Photo illustration of the four tasks* of task performed by a chronic stroke survivor (LHD, UEFM – 51). Left panel show s the two letter envelope tasks: folding the letter and inserting in an envelope. Right panel shows the two photo-album tasks: receiving the photo album and inserting photo in album sleeve.

The two letter-envelope subtasks were similar in that they involve manipulation of paper objects, which were lightweight and required stabilization. In contrast, the photo album was heavy and somewhat self-stabilizing, especially once received and placed on the table. Apart from these overt differences in task demands, the two letter-envelope tasks were also temporally contiguous with each other but were separated in time from the two photo-album tasks. In other words, “inserting the letter into an envelope” was always preceded by “folding the letter”. Similarly, “inserting a photo in an album sleeve” first required “receiving the album”, placing it on the table, opening it to a desired location (at the discretion of the participant). As will be noted later, this obvious separation of the letter-envelope task pair from the photo-album task pair reveals itself in the post-hoc analyses of the choice data.

### Outcome Measures

The primary outcome was the overall choice of motor strategy, which was quantified as the selection of one (unimanual, =0) or both hands (bimanual, =1) to accomplish the task goal. All participants were successful at accomplishing the goal of the four subtasks. For the subtasks for which manual roles are somewhat well-defined (fold letter, insert letter into envelope, insert photo in album sleeve), a chosen strategy was considered bimanual if both hands were engaged in task-relevant roles. Unlike these, there was a less clear differentiation of manual roles in the *Receive photo album* subtask, thus a chosen strategy was considered to be bimanual as long as the contralesional arm/hand came in contact with the object and/or provided assistance toward the successful completion of the goal.

The secondary outcome was the time taken to complete each of the four subtasks, or movement time. As tasks were performed in a relatively continuous manner, we implemented a discretization process to mark the start and end of each subtask (see Supplementary Material II). In general, start times were defined as when initial contact was made with the object of interest (letter or envelope or photo album), and end times were defined as when the goal was accomplished, e.g., when the last fold was completed, or letter was fully inserted or album came in contact with the table surface. Movement time was defined as the time elapsed between the start and end time points for each subtask.

Two evaluators blinded to the a-priori hypotheses of this study were trained to code the choice strategies as unimanual or bimanual and to discretize the four subtasks by identifying the start and end frames for movement time analysis.

### Statistical Analysis

All analyses were conducted using the R statistical computing package (version 3.5.1).

#### Primary analysis

Fisher’s Exact test was used to compare the proportion of bimanual choices between age-similar able-bodied controls and chronic stroke survivors.

In the subset of stroke survivors only, to assess the influence of the degree of motor impairment (UEFM) and the side of stroke lesion on bimanual choice (Strategy), we used nested mixed-effects multiple logistic regression. Below is the model form (in Wilkinson notation):

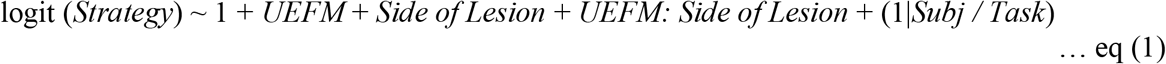

Random effects were modeled as *Task* nested within *Subject* (1| *Subj / Task*) because *Strategy* was repeatedly sampled over the four tasks within each subject. We compared this model to the null model as well as simpler reduced models using the Likelihood Ratio Tests (LRT). To systematically explore task-wise differences, we conducted a post-hoc analysis in which we separated the two letter-envelope tasks from the two photo-album tasks (n = 100, each) and repeated the nested mixed-effects model.

#### Secondary analysis

Independent two-sample Welch’s t-tests were used to compare average movement times between age-similar able-bodied controls and chronic stroke survivors across the four subtasks.

In the subset of stroke survivors only, we used mixed-effects multiple linear regression to test the influence of choice on movement time. Based on the previously studied influence of motor impairment on movement time (Chae et al. 2002; Kamper et al. 2002; Varghese and Winstein 2020), we included UEFM in the model as a covariate (form below):

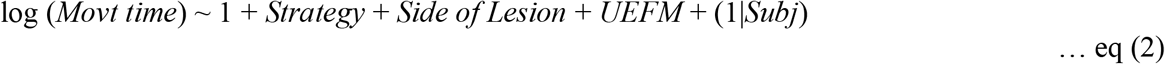

Consequently, we repeated the nested mixed-effects regression separately for each of the two pairs of tasks, i.e., letter-envelope tasks and photo-album tasks.

Other potential confounders (age, chronicity, and sex) were also tested for influence in the above models using a backward selection approach; those predictors that met a liberal cut-off of *p* = 0.2 were preserved in the final reduced model. Based on this selection process, none of the confounders met the cut-off p-value, except our hypothesized predictors. Continuous variables (age, chronicity, UEFM scores, and movement time) were assessed for normality. UEFM was converted to standardized z-scores. Distributions for chronicity and movement time were positively skewed and so they were log transformed. All necessary assumptions for Generalized Linear Models, including linearity, equality of variance, independence and normality of errors, and multicollinearity of independent variables, were tested and found adequate.

One-way ANOVA was used to compare age among the three groups, and Welch’s t-tests was used to compare chronicity and UEFM scores between LHD and RHD. Chi-square test was used to compare the proportion of females and males among the three groups. Significance level was set at p = 0.05.

## Results

Of the total 61 participants, 37 (60.6%) were male. Average age of the full sample was 60 years and of the stroke survivors was 59.7 years. Average chronicity was 5.4 years post-stroke and score on the UEFM was 42.2, indicating moderate impairment. Chronic stroke survivors consisted of equal numbers of individuals with left- (LHD) and right-hemisphere damage (RHD). RHD were on average slightly more chronic compared to those with LHD (difference of approximately 10 months); however, median chronicity was much more comparable between the two groups (difference of 2.73 months). Overall, there were no significant differences between the two groups with respect to age, chronicity or UEFM scores. Table 1 shows group-wise demographic information.

**Table 1.**
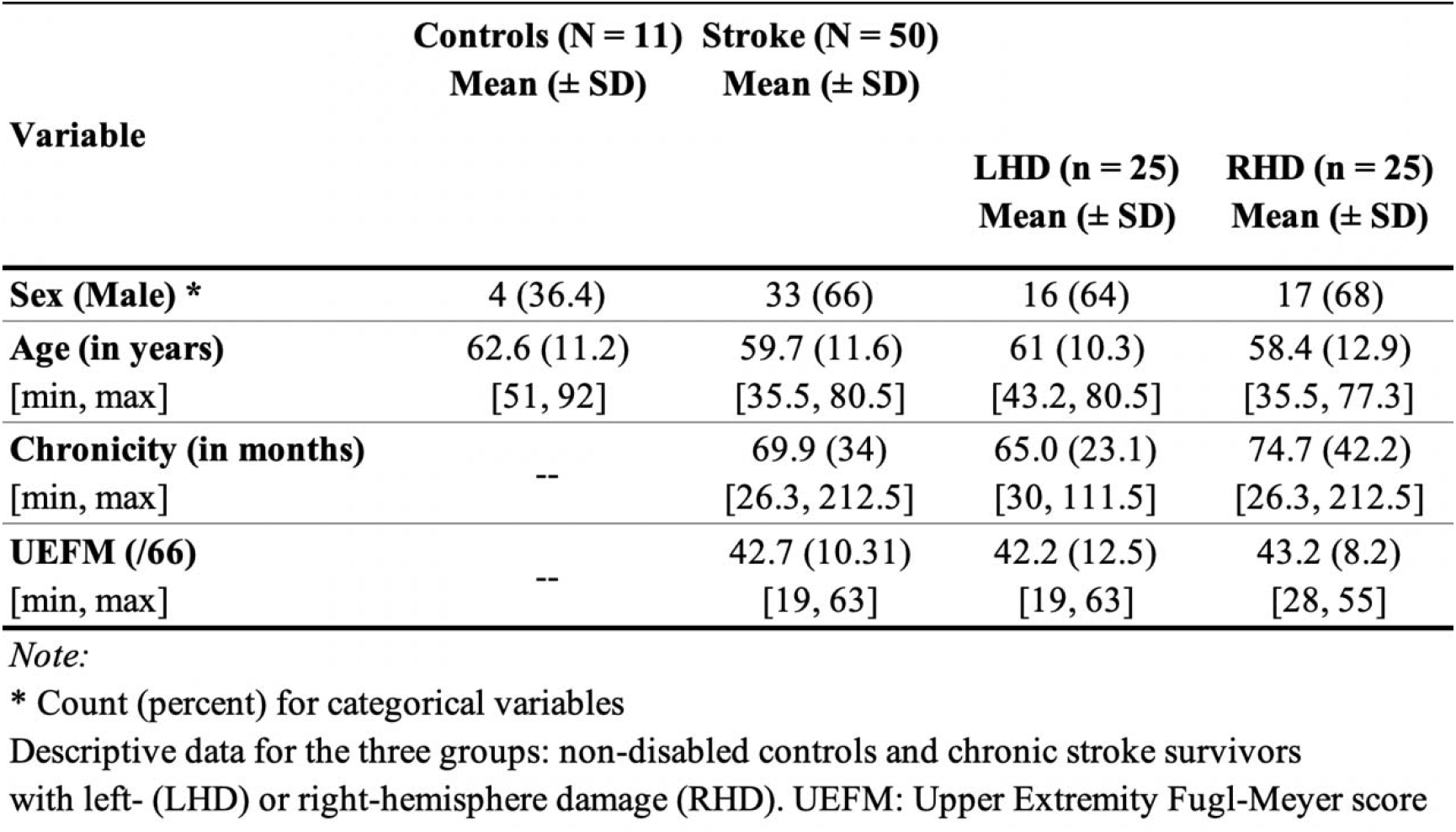
Participant Characteristics.

### The choice to use both hands is influenced by the degree of motor impairment and the side of lesion

Compared to age-similar able-bodied adults who exclusively chose a bimanual strategy, chronic stroke survivors were significantly less likely to choose both hands for all tasks (Fisher’s Exact *p* < 0.0001). Of the 200 choices, 131 (65.5%) were bimanual.

The final model (eq. 1) was significantly different from a null model (LRT χ^2^ (4) = 25.55, *p* = 1.18e-5) and from a model with side of lesion alone (LRT χ^2^ (2) = 23.64, *p* = 7.35e-6), and was only modestly different from a model with UEFM alone (LRT χ^2^ (2) = 4.82, *p*= 0.09).

There was a significant effect of motor impairment on choice, such that those with a higher UEFM scores (less severe) were more likely to choose both hands together (*p*= 7.77e-05). After taking into account the effect of motor impairment, there was also a significant, albeit small, effect of the side of lesion (*p* = 0.033) such that those with left hemisphere damage (LHD) were more likely to use both hands together compared to those with right hemisphere damage (RHD). Table 2 (column A) shows standardized estimates and model performance measures from the mixed-effects logistic regression.

**Table 2.** Standardized coefficients ± SE from nested mixed-effects logistic regression.

|  | (A)<br>Overall | (B)<br>Letter-Envelope tasks | (C)<br>Photo-Album tasks |
| --- | --- | --- | --- |
| (Intercept) | 1.251 ± 0.31 *** | 13.569 ± 2.89 *** | 0.686 ± 0.43 *** |
| UEFM <sup>§</sup> | 1.002 ± 0.25 *** | 0.654 ± 1.91 | 1.36 ± 0.43 ** |
| Side of Lesion <sup>#</sup> | -0.838 ± 0.39 * | 29.22 ± 11.15 ** | -1.441 ± 0.61 * |
| UEFM × Side of Lesion | -0.047 ± 0.42 | 42.67 ± 11.50 *** | -0.498 ± 0.64 |
| Task : Subj Intercept (sd) | 0.0007 | 71.82 | 0.0001 |
| Subj. Intercept (sd) | 0.505 | 23.34 | 0.77 |
| N | 200 | 100 | 100 |
| Log Likelihood | -110.89 | -22.12 | -55.22 |
| AIC | 233.78 | 56.24 | 122.45 |

\*\*\* $p < 0.001$ ; \*\* $p < 0.01$ ; \* $p < 0.05$
§ standardized UEFM

### The choice to use both hands is influenced by a task-specific interaction between motor impairment and side of lesion

There were systematic differences in the proportion of bimanual choices such that the two letter-envelope tasks—folding the letter (80%) and inserting the letter into the envelope (86%)—were nearly twice as likely to elicit a bimanual choice in stroke survivors compared to the two photo-album tasks—receiving album (44%) and inserting photo in album sleeve (52%).

Results of task-wise mixed models revealed stark differences between the two task pairs with respect to the effect of motor impairment and side of lesion on bimanual choice. Specifically, for the letter-envelope task pair, there was no main effect of UEFM but a strong interaction effect (β = 42.67, *p* = 2.06e-04) suggesting that the probability of choosing a bimanual strategy rises steeply with UEFM for those with right hemisphere damage (RHD), but does not significantly vary in those with LHD. Moreover, the intercept (at the mean UEFM, 42.7) was also higher in RHD compared to LHD (β = 29.22, *p* = 0.008) suggesting the opposite direction of effect from the overall model. That is, for the letter-envelope tasks, those with RHD were more likely to choose both hands together compared to those with LHD. Results for the photo-album tasks were consistent with the overall effects observed for all four tasks. That is, those with LHD are more likely to choose a bimanual strategy for the photo-album tasks. Table 2 (columns B & C) show standardized estimates from the separate task-wise models.

To visualize these results, we used estimates from the logistic regression to compute and plot predicted probabilities of using a bimanual strategy. Fig. 2 shows raw data and model-fitted probabilities for LHD and RHD across UEFM scores.

**Figure 2.**
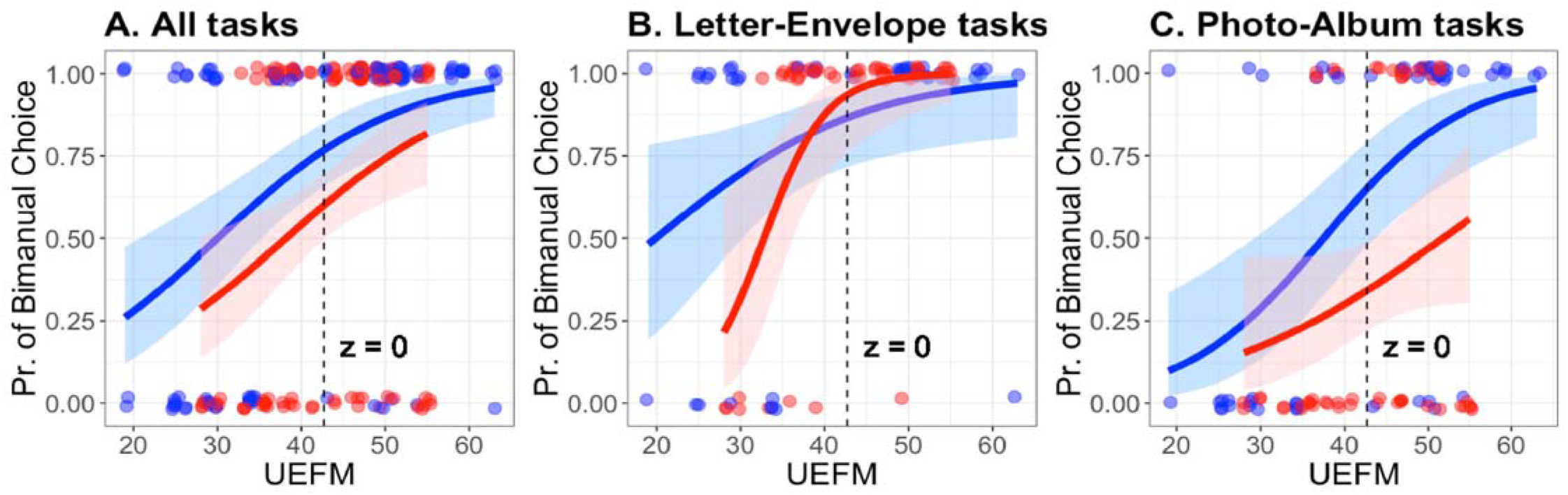
Raw data (1 = bimanual, 0 = unimanual) and model-fitted probabilities (Pr.) of bimanual choice for left- (LHD, blue) and right-hemisphere damage (RHD, red) groups. Logistic model fits A. across all four tasks (n = 200), B. letter-envelope tasks (n = 100), and C. photo-album tasks (n = 100). Dashed line corresponds to the mean UEFM score (z = 0) and the intercept of the model fit.

### Movement time was significantly longer in chronic stroke survivors but not different between those who chose a unimanual compared to a bimanual strategy

Compared to able-bodied adults, chronic stroke survivors took 2.4 times longer on average across tasks (*t* = 7.75, *p* <0.0001); 1.5 times longer for the letter-envelope tasks (*t* = 7.96, *p* <0.0001), and 2.5 times longer for the photo-album tasks (*t* = 5.54, *p* <0.0001). After taking into account the degree of motor impairment and the side of lesion, there were, no differences between individuals who chose a unimanual compared to a bimanual strategy (Table 3)

**Table 3.** Average movement times for stroke and able-bodied control participants. Standardized coefficients from nested mixed-effects linear regression in stroke survivors only.

|  |  | (A)<br>Overall | (B)<br>Letter-Envelope | (C)<br>Photo-Album |
| --- | --- | --- | --- | --- |
| **Movt. Time**<br>(mean ± SD, sec) | **Stroke** | 10.9 ± 9.2 | 16.1 ± 5.9 | 5.8 ± 8.9 |
| | **Control** | 4.5 ± 3.4 | 5.8 ± 3.5 | 2.3 ± 1 |
| (Intercept) | | 1.467 ± 0.14 | 2.674 ± 0.15 | 1.189 ± 0.17 |
| **Strategy** | | 0.683 ± 0.15 | -0.112 ± 0.15 | 0.239 ± 0.20 |
| **Side of Lesion** <sup>#</sup> | | 0.152 ± 0.13 | 0.085 ± 0.11 | 0.108 ± 0.18 |
| **UEFM** <sup>§</sup> | | -0.266 ± 0.07 | -0.20 ± 0.06 | -0.115 ± 0.09 |
| **Subj. Intercept (sd)** |  | 0 | 0.084 | 0 |
| **Obs. Residual (sd)** |  | 0.92 | 0.52 | 0.88 |
| **N** |  | 200 | 100 | 100 |
| **Log Likelihood** |  | -270.77 | -82.83 | -131.31 |
| **AIC** |  | 553.54 | 177.67 | 274.62 |

\*\*\* $p < 0.001$ ; \*\* $p < 0.01$ ; \* $p < 0.05$
§ standardized UEFM

Across all participants, the letter-envelope tasks took significantly longer (14.4 s) compared to the photo-album tasks (5.2 s). Figure 3 displays boxplots for log-transformed movement time comparisons between strategy choices for stroke groups and controls.

**Figure 3.**
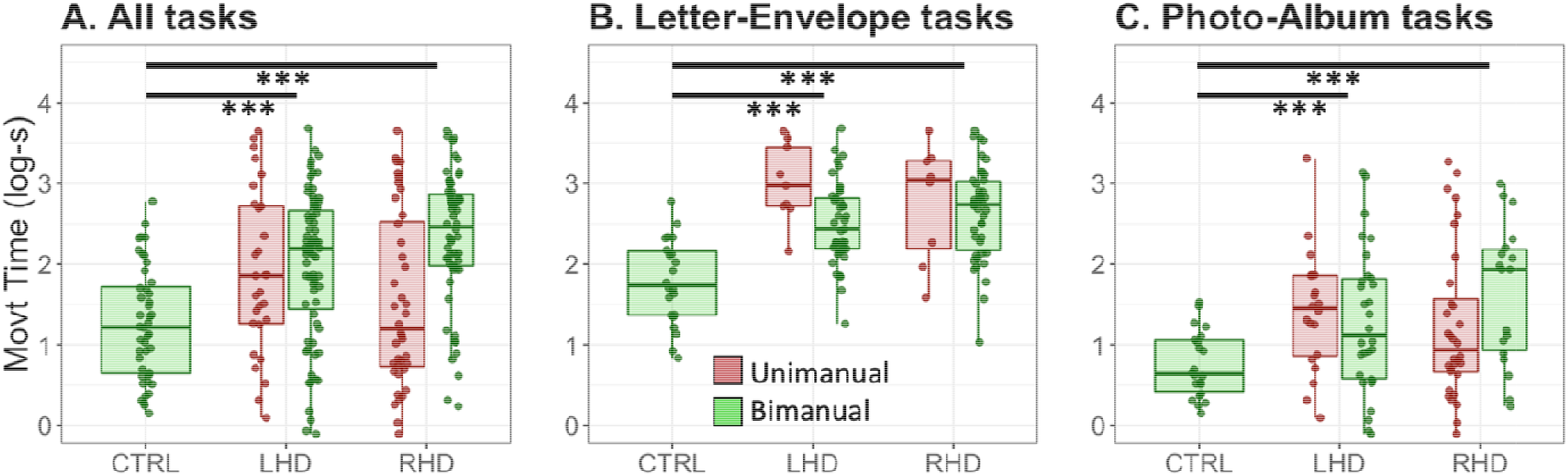
Average log-transformed movement times A. across all four tasks (n = 200), B. letter-envelope tasks (n = 100), and C. photo-album tasks (n = 100). Non-disabled control participants (CTRL) were significantly faster compared to chronic stroke survivors who chose a bimanual strategy. As predicted, there were no differences in movement time between those stroke survivors who chose a unimanual compared to a bimanual strategy. ***p< 0.001

## Discussion

A goal of rehabilitation after stroke is to promote pre-stroke levels of (paretic) arm use for everyday functional activities—many of which require the use of both hands. The central question in this study was: when faced with tasks that demand the use of both hands, how do chronic stroke survivors choose to solve them; what factors determine whether the paretic arm is engaged as part of the solution? This is the first study to investigate self-selected and task-specific choices made by chronic stroke survivors in the context of bimanual tasks.

By covertly observing spontaneous use behaviors for two distinct types of bimanual tasks, we found that, compared to age-similar, able-bodied adults, chronic stroke survivors were significantly less likely to spontaneously choose a bimanual strategy. Generally consistent with previous reports, (Rinehart et al. 2009; Thrane et al. 2011; Bailey et al. 2014; Yadav et al. 2019) the choice to engage both hands together depended on the degree of impairment and the side of stroke lesion. Notably, we extended prior observations to show that the influence of these factors on choice varied based on task demands. This is novel because arm use metrics in those previous studies (Sterr et al. 2002; Vega-González and Granat 2005; Michielsen et al. 2012; Bailey et al. 2014, 2015) were often collapsed across various tasks, and so, task-wise effects, if any, may have been lost.

### Bimanual use emerges from an interaction between task demands and lateralized motor control processes

As alluded to in the Introduction to this study, we expected that the choice of strategy would emerge from an interaction of task demands with lateralized motor control processes. For the letter-envelope task, we suspected greater engagement of the left hand due to stabilization requirements, and found that the likelihood of choosing both hands together rose sharply for those with moderate impairment (UEFM > 30) of the left hand (i.e., RHD), but not the right (i.e., LHD). Conversely, for the self-stabilizing photo-album, we expected greater engagement of the right hand due to strength and precision requirements and found that the probability of a bimanual strategy was greater for those with LHD compared to RHD, regardless of the degree of motor impairment.

These task-specific effects seem consistent with the predictions of a theoretical framework known as the dynamic dominance hypothesis (DDH). The traditional view of limb dominance attributes unimanual preferences for the right hand to its general superiority for motor skills while the left hand is regarded as a weaker counterpart. Conversely, the DDH proposes that each limb is proficient for a different aspect of task performance; the left hand for position stabilization and the right hand for predictive control of reach. In the context of a bimanual task, the two hands are preferentially selected by the nervous system to assume roles consistent with their proficient controller.

In able-bodied adults, Stone et al (Stone et al. 2013) support the idea that these spontaneously preferred roles are dissociable between limbs. They observed that for a 3D model building task, even when stabilization requirements were met within the task and the left hand was free to reach and retrieve, it did not do so with any greater frequency than when stabilization was required. Instead, the left hand rested or “hovered”, seemingly unable to disengage from its stabilization role. Recently, Woytowicz and colleagues (Woytowicz et al. 2018) reported that switching preferred roles of the left and right hand during a simulated “bread-cutting” task was in fact non-optimal for motor performance. By examining relevant characteristics of the movement, they demonstrated that the right hand showed straighter reaching trajectories but was poor at stabilizing, whereas the left hand exhibited more stable holding performance, but was poor at reaching.

Taken together, it appears that selecting both hands to solve a bimanual task problem emerges from an interaction between task demands and specialized control of each limb, rather than global dominance. Our findings in chronic stroke survivors suggest that this task-specific engagement of each limb persists after a stroke, especially when there is sufficient capacity in the paretic hand.

### A capacity threshold for bimanual use

The preceding discussion raises another question: what amounts to sufficient motor capacity? The relationship between use and sensorimotor capacity has been previously explained through the threshold hypothesis (Schweighofer et al. 2009). This hypothesis suggests that when contralesional arm motor capacity exceeds a certain “functional threshold,” there is a sharp rise in the likelihood of its use. Although highly variable, the predicted probabilities from our logistic model render some support for this hypothesis. In the bimanual context, such a threshold likely varies from task to task and may differ between those with left-versus right-hemisphere damage. This is visible on Figure 2 where it appears that for the RHD group, the UEFM score at which a switch from unimanual to bimanual choice occurs is more distinct for the letter-envelope tasks (UEFM ~ 30) compared to the photo-album tasks. Conversely, for the LHD group, the threshold is more discernible for the photo-album tasks (UEFM ~ 35-40).

In addition to these task- and side of lesion-wise trends in capacity threshold, it seems interesting that compared to previous studies (Thrane et al. 2011; Han et al. 2013; Yadav et al. 2019), including our recent work (Buxbaum et al. 2019), which reported paretic limb non-use as a particular issue in those with mild-to-moderate impairment (UEFM 35-45), the capacity threshold may be lower for bimanual tasks, suggesting that bimanual tasks impose greater demands on the neuromotor system thereby compelling the engagement of the paretic limb. Nonetheless, as recently reported by Yadav et al (Yadav et al. 2019), bilateral use is diminished significantly for those with most severe impairment regardless of side of lesion.

### The influence of experience, habits, and perception on spontaneous bimanual use

A final but crucial perspective for interpreting these results is that, through a unique covert observation paradigm, we have been able to capture spontaneous use reflected in self-selected choices. In able-bodied adults, the spontaneous choice to use both hands appears to be a well-established behavioral response that reflects a complex implicit decision-making process. Studies in able-bodied and disabled adults demonstrate that this decision is driven by prior habits and experiences (Han et al. 2013; Kim et al. 2018), instantaneous negotiation of the salient and non-salient features of the task and environment (e.g., task goals, object affordance) (Mamolo et al. 2004, 2006; Stone et al. 2013), and actual (estimated) and perceived consequences of a given action, including its associated costs (e.g., time or energetic costs) and likelihood of success. (Witt et al. 2004; Shadmehr et al. 2010, 2016; Schweighofer et al. 2015)

Stroke survivors (both LHD and RHD) in this study were approximately 5 years post-stroke—a period long enough to acquire new experiences and habits (such as learning new skills with the ipsilesional left hand in LHD) or even reinforce pre-stroke habits (such as continued success in using the ipsilesional right hand in RHD after initial failures using the paretic left hand) (see Jones 2017 for a review). Given these considerations, spontaneous choice behaviors observed in our sample of stroke survivors are likely to be largely stereotypical and successful compensatory behaviors—behaviors that the individual may have implicitly deemed useful toward attainment of every day bimanual task goals. With regards to costs associated with selection of a motor strategy, we did not observe any differences in movement time between stroke survivors who chose a unimanual compared to a bimanual strategy, lending support to the idea that individuals would be likely to choose a motor strategy that represents the most efficient strategy in terms of time to complete the task (but see an important limitation below).

### Limitations

Interpretations of these findings are limited by several limitations. The retrospective design of this study and the relatively small sample size limit the generalization of these observations. Nonetheless, the observational nature of our analysis provides an important and unique perspective of ecologically valid and task-specific arm choice after stroke. Prospective experiments are needed to assess bimanual use by systematically varying task demands to test the interaction between task demands and lateralized motor control. Another limitation was that the severity of motor impairment was restricted in our cohort of participants with the minimum UEFM score being 19 in LHD and 28 in RHD. To systematically assess if capacity threshold for bimanual use varies by the type of task and side of lesion, future studies should consider a wider range of motor impairment scores. Lastly, we are cautious in interpreting the results of our secondary analysis due to the unequal sample sizes for each strategy. A within-subject design in which each participant performs the task unimanually and bimanually would be more appropriate to accurately test this hypothesis. The typical forced condition of the AAUT did not provide for the unimanual corollary required for this type of comparison.

## Conclusion

In conclusion, the present study provides preliminary evidence for self-selected and task-specific choice for ecologically valid bimanual tasks in chronic stroke survivors. Unlike age-similar able-bodied adults, chronic stroke survivors *do not* spontaneously choose both hands to solve routine bimanual tasks. The probability of choosing both hands increases when the contralesional arm is less impaired. Importantly, the effect of motor impairment is modified both by the side of lesion and the type of task. We argue that our findings seem inconsistent with the predictions of a traditional global dominance model. Instead, in chronic stroke survivors, bimanual use emerges from a task-specific interaction between motor impairment and the side of lesion, such that when there is sufficient motor capacity, the paretic hand is preferentially selected by the nervous system to assume a role consistent with its specialized controller.

## Data Availability

The data table and code for analysis are available on the first author’s OSF repository.

https://doi.org/10.17605/osf.io/uh574

## Acknowledgements

Dr. Laurel Buxbaum (Moss Rehabilitation Research Institute) for contributing their dataset to this analysis. Drs. James Gordon (USC) and Robert Sainburg (Pennsylvania State University) for helpful discussions. Cassandra Castillo and Antonio Raymundo, students of the Engineering for Health Academy program, Francisco Bravo Medical Magnet High School, LAUSD, for coding the video data.

## Data and Code Availability

The data table and code for analysis are available on the first author’s OSF repository: https://doi.org/10.17605/osf.io/uh574

## References

Bailey RR, Klaesner JW, Lang CE (2015) Quantifying Real-World Upper-Limb Activity in Nondisabled Adults and Adults with Chronic Stroke. Neurorehabil Neural Repair 29:969–978. https://doi.org/10.1177/1545968315583720

Bailey RR, Klaesner JW, Lang CE (2014) An accelerometry-based methodology for assessment of real-world bilateral upper extremity activity. PLoS One 9:. https://doi.org/10.1371/journal.pone.0103135

Buxbaum LJ, Varghese R, Stoll H, Winstein CJ (2020) Predictors of arm non-use in chronic stroke: a preliminary investigation. Neurorehabil Neural Repair (in press). Available as preprint: https://doi.org/10.1101/702159

Chae J, Yang G, Park BK, Labatia I (2002) Delay in initiation and termination of muscle contraction, motor impairment, and physical disability in upper limb hemiparesis. Muscle and Nerve 25:568–575. https://doi.org/10.1002/mus.10061

Coelho CJ, Przybyla A, Yadav V, Sainburg RL (2013) Hemispheric differences in the control of limb dynamics: A link between arm performance asymmetries and arm selection patterns. J Neurophysiol 109:825–838. https://doi.org/10.1152/jn.00885.2012

Franck JA, Smeets RJEM, Seelen HAM (2019) Changes in actual arm-hand use in stroke patients during and after clinical rehabilitation involving a well-defined arm-hand rehabilitation program: A prospective cohort study. PLoS One 14:. https://doi.org/10.1371/journal.pone.0214651

Fugl Meyer AR, Jaasko L, Leyman I (1975) The post stroke hemiplegic patient. I. A method for evaluation of physical performance. Scand J Rehabil Med

Guiard Y (1987) Asymmetric division of labor in human skilled bimanual action: The kinematic chain as a model. J Mot Behav 19:486–517. https://doi.org/10.1080/00222895.1987.10735426

Haaland KY, Mutha PK, Rinehart JK, et al (2012) Relationship between arm usage and instrumental activities of daily living after unilateral stroke. Arch Phys Med Rehabil 93:1957–1962. https://doi.org/10.1016/j.apmr.2012.05.011

Han CE, Kim S, Chen S, et al (2013) Quantifying arm nonuse in individuals poststroke. Neurorehabil Neural Repair 27:439–447. https://doi.org/10.1177/1545968312471904

Jones TA (2017) Motor compensation and its effects on neural reorganization after stroke. Nat. Rev. Neurosci. 18: 267–280

Kamper DG, McKenna-Cole AN, Kahn LE, Reinkensmeyer DJ (2002) Alterations in reaching after stroke and their relation to movement direction and impairment severity. Arch Phys Med Rehabil 83:702–707. https://doi.org/10.1053/apmr.2002.32446

Kilbreath SL, Heard RC (2005) Frequency of hand use in healthy older persons. Aust J Physiother 51:119–122. https://doi.org/10.1016/S0004-9514(05)70040-4

Kim S, Park H, Han CE, et al (2018) Measuring habitual arm use post-stroke with a bilateral time-constrained reaching task. Front Neurol 9:1–5. https://doi.org/10.3389/fneur.2018.00883

Mamolo CM, Roy E a, Rohr LE, Bryden PJ (2006) Reaching patterns across working space: the effects of handedness, task demands, and comfort levels. Laterality 11:465–492. https://doi.org/10.1080/13576500600775692

Mamolo CM, Roy EA, Bryden PJ, Rohr LE (2004) The effects of skill demands and object position on the distribution of preferred hand reaches. Brain Cogn 55:349–351. https://doi.org/10.1016/j.bandc.2004.02.041

Michielsen ME, Selles RW, Stam HJ, et al (2012) Quantifying nonuse in chronic stroke patients: A study into paretic, nonparetic, and bimanual upper-limb use in daily life. Arch Phys Med Rehabil 93:1975–1981. https://doi.org/10.1016/j.apmr.2012.03.016

Przybyla A, Coelho CJ, Akpinar S, et al (2013) Sensorimotor performance asymmetries predict hand selection. Neuroscience. https://doi.org/10.1016/j.neuroscience.2012.10.046

Rand D, Eng JJ (2015) Predicting daily use of the affected upper extremity 1 year after stroke. J Stroke Cerebrovasc Dis 24:274–283. https://doi.org/10.1016/jjstrokecerebrovasdis.2014.07.039

Rinehart JK, Singleton RD, Adair JC, et al (2009) Arm use after left or right hemiparesis is influenced by hand preference. Stroke 40:545–550. https://doi.org/10.1161/STROKEAHA.108.528497

Sainburg RL (2002) Evidence for a dynamic-dominance hypothesis of handedness. Exp Brain Res 142:241–258. https://doi.org/10.1007/s00221-001-0913-8

Schweighofer N, Han CE, Wolf SL, et al (2009) A Functional Threshold for Long-Term Use of Hand and Arm Function Can Be Determined: Predictions From a Computational Model and Supporting Data From the Extremity Constraint-Induced Therapy Evaluation (EXCITE) Trial. Phys Ther. https://doi.org/10.2522/ptj.20080402

Schweighofer N, Xiao Y, Kim S, et al (2015) Effort, success, and nonuse determine arm choice. J Neurophysiol 114:551–559. https://doi.org/10.1152/jn.00593.2014

Shadmehr R, De Xivry JJO, Xu-Wilson M, Shih TY (2010) Temporal discounting of reward and the cost of time in motor control. J Neurosci 30:10507–10516. https://doi.org/10.1523/JNEUROSCI.134310.2010

Shadmehr R, Huang HJ, Ahmed AA (2016) A Representation of Effort in Decision-Making and Motor Control. Curr Biol 26:1929–1934. https://doi.org/10.1016/j.cub.2016.05.065

Sterr A, Freivogel S, Schmalohr D (2002) Neurobehavioral aspects of recovery: Assessment of the learned nonuse phenomenon in hemiparetic adolescents. Arch Phys Med Rehabil. https://doi.org/10.1053/apmr.2002.35660

Stone KD, Bryant DC, Gonzalez CLR (2013) Hand use for grasping in a bimanual task: Evidence for different roles? Exp Brain Res 224:455–467. https://doi.org/10.1007/s00221-012-3325-z

Taub E, Crago JE, Uswatte G (1998) Constraint-induced movement therapy: A new approach to treatment in physical rehabilitation. Rehabil Psychol 43:152–170. https://doi.org/10.1037/0090-5550.43.2.152

Thrane G, Emaus N, Askim T, Anke A (2011) Arm use in patients with subacute stroke monitored by accelerometry: Association with motor impairment and influence on self-dependence. J Rehabil Med 43:299–304. https://doi.org/10.2340/16501977-0676

Uswatte G, Giuliani C, Winstein C, et al (2006) Validity of Accelerometry for Monitoring Real-World Arm Activity in Patients With Subacute Stroke: Evidence From the Extremity Constraint-Induced Therapy Evaluation Trial. Arch Phys Med Rehabil 87:1340–1345. https://doi.org/10.1016/j.apmr.2006.06.006.

Varghese R, Winstein CJ (2020) Relationship Between Motor Capacity of the Contralesional and Ipsilesional Hand Depends on the Side of Stroke in Chronic Stroke Survivors With Mild-to-Moderate Impairment. Front. Neurol. 10:1340. https://doi.org/10.3389/fneur.2019.01340

Vega-González A, Granat MH (2005) Continuous monitoring of upper-limb activity in a free-living environment. Arch Phys Med Rehabil 86:541–548. https://doi.org/10.1016/j.apmr.2004.04.049

Winstein C, Kim B, Kim S, et al (2019) Dosage Matters: A Phase IIb Randomized Controlled Trial of Motor Therapy in the Chronic Phase after Stroke. Stroke 50:1831–1837. https://doi.org/10.1161/STROKEAHA.118.023603

Witt JK, Proffitt DR, Epstein W (2004) Perceiving distance: A role of effort and intent. Perception. https://doi.org/10.1068/p5090

Woytowicz EJ, Westlake KP, Whitall J, Sainburg RL (2018) Handedness results from complementary hemispheric dominance, not global hemispheric dominance: evidence from mechanically coupled bilateral movements. J Neurophysiol 120:729–740. https://doi.org/10.1152/jn.00878.2017

Yadav G, Haaland KY, Mutha PK (2019) Laterality of Damage Influences the Relationship between Impairment and Arm Use after Stroke. J Int Neuropsychol Soc 25:470–478. https://doi.org/10.1017/S1355617718001261

